# Baseline Peripheral Blood Counts and Outcomes in Patients Presenting with COVID-19

**DOI:** 10.1101/2022.06.10.22276256

**Authors:** Preethi Jeyaraman, Pronamee Borah, Omender Singh, Arun Dewan, Nitin Dayal, Rahul Naithani

**Affiliations:** Hematology and Bone Marrow Transplant Division, Max Superspecialty Hospital, Saket, New Delhi; Department of Critical Care Medicine, Max Superspecialty Hospital, Saket, New Delhi; Department of Internal Medicine, Max Smart Superspeciality Hospital, Saket, New Delhi; Department of Lab Medicine, Max Superspeciality Hospital, Saket, New Delhi

**Author notes:** **Corresponding Author** Dr Rahul Naithani, Director -Hematology and Bone Marrow Transplant Division, Max Superspecialty Hospital, Saket, New Delhi, Ph no, Email id. Financial disclosure: No funding was taken for conducting this study.

**Keywords:** Covid-19, blood counts, neutrophil lymphocyte ratio, eosinopenia

## Abstract

**Background:** SARS-CoV-2 pandemic has significant impact on hematopoietic system.

**Objective:** To report the incidence and pattern of baseline hematological parameters in patients with COVID-19 and their association with severity of disease and outcome.

**Methods:** Retrospective observational study.

**Results:** A total of 440 patients were included in the study. The mean age of the study cohort was 47.5 ±15.8 years. Fifty percent of patients had at least 1 comorbidity. ICU stay was required in 125 (39.6%) patients. Overall mortality in the study cohort was 3.52%. The average age of patients who died was significantly higher than that of patients who were alive (65.1 years vs 46.5 years; p= 0.000). DM, HTN, CAD and CKD were all associated with higher incidence of ICU stay and mortality. Lymphopenia < 1×10^9^/μl was observed in 24.3% and eosinopenia was noted in 44.3% patients. Leukocytosis>11×10^9^/μl was seen in 8.2 % of patients. The median neutrophil lymphocyte ratio (NLR) of whole cohort was 2.63. NLR, Lymphopenia, eosinopenia, leucocytosis, D dimer, lactate dehydrogenase (LDH), ferritin and IL6 levels all were associated with need for ICU transfer and mortality. Hemoglobin, red cell distribution width (RDW), PT and aPTT correlated with need for ICU transfer but not with mortality. Ferritin cutoff ≥751 ng/ml and IL6 levels ≥64pg/ml was able to identify all deaths. Ferritin (0.989) and IL-6 (0.985) had very high negative predictive value.

**Conclusions:** Peripheral blood counts at time of hospitalization is a simple tool to predict outcomes in patients admitted with Covid-19.

## Introduction

Severe acute respiratory syndrome coronavirus 2 (SARS-CoV-2) causing coronavirus disease 2019 (COVID-19) with origin in Wuhan, China has caused a pandemic[1]. It is now considered a systemic disease with multiple organ manifestations including cardiovascular, respiratory, gastrointestinal and neurological being reported[2]. Even then, literature on hematological manifestations is scarce.

COVID-19 can have significant impact on the haematological system in the form of cytopenia and disseminated intravascular coagulation[3,4]. Tan et al have proposed a model based on lymphocyte counts at two time points; patients with less than 20% lymphocytes at days 10-12 from the onset of symptoms and less than 5% at days 17-19 have the worst prognosis[5].

Thrombocytopenia has been associated with severe disease. Increased platelet lymphocyte ratio and peak in platelet count have been reported to be associated with prolonged hospitalisation. Coagulation abnormalities are found commonly in COVID-19 patients[6]. Criteria for disseminated intravascular coagulation (DIC) was fulfilled in 71.4% of non-survivors as compared to 0.6% of survivors. The median time from admission to DIC manifestation was 4 days (range: 1-12 days).

One study correlated presence of lymphopenia to severity of disease[7] while another found neutrophilia to be associated with risk of mortality[8]. Role of monocytes in inflammation of Covid-19 has been recently published[9]. Pakos et al have demonstrated association of lower monocyte count, platelet count and higher neutrophil-to-lymphocyte ratio (NLR) and mortality[10].

We report the incidence and pattern of baseline hematological parameters in patients with COVID-19 presenting to our center and their association with severity of disease.

## Methods

This is a retrospective observational study conducted at tertiary care teaching hospital. Patients >18 years of age and who were admitted with COVID-19 infection from 10-31 May 2020 were enrolled in the study. A diagnosis of severe acute respiratory syndrome coronavirus 2 (SARS-CoV-2)-infection was based on quantitative real-time reverse transcriptase-polymerase chain reaction (qRT-PCR) of nasal and/and or pharyngeal swabs. The baseline demographic data and haematological parameters (at the time of hospitalisation) like complete blood count, differential leukocyte count and coagulation abnormalities were extracted from electronic health records and entered in Microsoft Excel spreadsheet. The nadir and peak haematological parameters were recorded in patients in whom these data were available. All patients received various treatments as per physician discretion, institutional protocol in accordance with the national guidelines for the management of COVID-19, issued by the government from time to time[11]. A variety of medicines were used in these patients and included hydroxychloroquine (HCQ), azithromycin, doxycycline, ivermectin, remdesivir, other broad spectrum antibiotics, steroids (methylprednisolone or dexamethasone), tocilizumab and oxygen support/ventilation as required[12]. Criteria for classifying patients into severe category were as per the clinical management protocols of government of India[11]. Outcomes including ICU transfer and death were recorded. Study was approved by institutional review board.

### Statistical analysis

Data was described in percentages for categorical variables and as the mean ± standard deviation values in case of continuous variables. For categorical data, comparisons were made by using the Chi square**/**Fisher exact test, for quantitative data by *t* test/F-test and for non-normally distributed quantitative variables by the Mann-Whitney/Kruskal Wallis test. Associations between severity and outcomes of haematological parameters was done with Pearsons correlation coefficient and spearman correlation coefficient.

Data was analysed with SPSS v 23 software. *P* value ≤0.05 was considered significant in all statistical evaluations.

## Results

A total of 440 patients admitted during the study period were included in the study. Demographic characteristics of patients included in the study have been summarized in Table1. The mean age of the study cohort was 47.5 ±15.8 years. Males predominated the cohort (67%). Fifty percent of patients had at least 1 comorbidity which included diabetes mellitus (DM) (32%), hypertension (HTN) (35%), coronary artery disease (CAD) (8.6%), chronic kidney disease (CKD) (5%), obstructive airway disease(3.9%) and malignancy(1.8%). Eight patients were on immunosuppressive medication. Mean duration of hospital stay was 11.4±4.5 days. ICU stay was required in 125 (39.6%) patients. Overall mortality in the study cohort was 3.52%. The average age of patients having severe disease, requiring ICU admission was higher (p=0.000). Male patients were prone for severe disease (p=0.007) and ICU stay (p= 0.001). The average age of patients who died was significantly higher than that of patients who were alive (65.1 years vs 46.5 years; p= 0.000). DM, HTN, CAD and CKD were all associated with higher incidence of ICU stay and mortality. Presence of malignancy or immunosuppressive medication did not correlate with ICU stay and mortality. Severe disease and ICU stay was more in patients with any comorbidity as compared with those who did not have.

**Table 1.** Demographic characteristics of cohort

|  | Total | ICU admissions |  |  | Mortality |  |  |
| --- | --- | --- | --- | --- | --- | --- | --- |
|  | N= 440 | Yes<br>125 | No<br>315 | P value | Yes<br>15 | No<br>425 | P value |
| Age (Years) | 47.5 ± 15.8<br>(18 to 95) | 56.6 ±12.8 | 43.9 ±15.4 | p=0.000 | 65.1±12.8 | 46.9±15.5 | 0.000 |
| Male | 295(67%) | 98 | 197 | 0.001 | 11 | 284 | 0.782 |
| Female | 145(33%) | 27 | 118 |  | 4 | 141 |  |
| Comorbidity |  |  |  |  |  |  |  |
| Any | 220 (50%) | 103 | 117 | 0.000 | 14 | 206 | 0.001 |
| Diabetes | 141 (32%) | 70 | 71 | 0.000 | 8 | 133 | 0.07 |
| Hypertension | 154 (35%) | 78 | 76 | 0.000 | 11 | 143 | 0.004 |
| CAD | 38 (8.6%) | 24 | 14 | 0.000 | 6 | 32 | 0.00 |
| CKD | 22 (5%) | 19 | 3 | 0.000 | 3 | 19 | 0.033 |
| COPD | 17 (3.9%) | 9 | 8 | 0.029 | 1 | 16 | 0.452 |
| Cancer | 8 (1.8%) | 3 | 5 | 0.693 | 1 | 7 | 0.244 |
| Immunosuppressant | 8 (1.8%) | 4 | 4 | 0.232 | 0 | 8 | 1.00 |
| Hospital stay (Days) | 11.4 ± 4.4 | 14.0±5.2 | 10.4±3.6 | 0.000 | 13.3±8.1 | 11.3± 4.2 | 0.15 |

CAD-Coronary artery disease; CKD- chronic kidney disease, COPD-chronic obstructive pulmonary disease

Table 2 summarises the haematological parameters at admission. Thrombocytopenia < 1 lakh was seen in only 1 patient (0.2%). Lymphopenia < 1×10^9^/μlwas observed in 24.3% and eosinopenia was noted in 44.3% patients. Leukocytosis>11×10^9^/μlwas seen in 8.2 % of patients. The median neutrophil lymphocyte ratio (NLR) of whole cohort was 2.63. Total leukocyte count (TLC), absolute neutrophil count (ANC), absolute eosinophil count (AEC), absolute lymphocyte count (ALC) and NLR correlated with need for ICU transfer and mortality. Lymphopenia, eosinopenia, leucocytosis, D dimer, lactate dehydrogenase (LDH), ferritin and IL6 levels all were associated with need for ICU transfer and mortality. Prolonged PT was seen in 60.5% and prolonged aPTT was seen in 10.5% of patients. Hemoglobin, red cell distribution width (RDW), PT and aPTT correlated with need for ICU transfer but not with mortality.

**Table 2.**
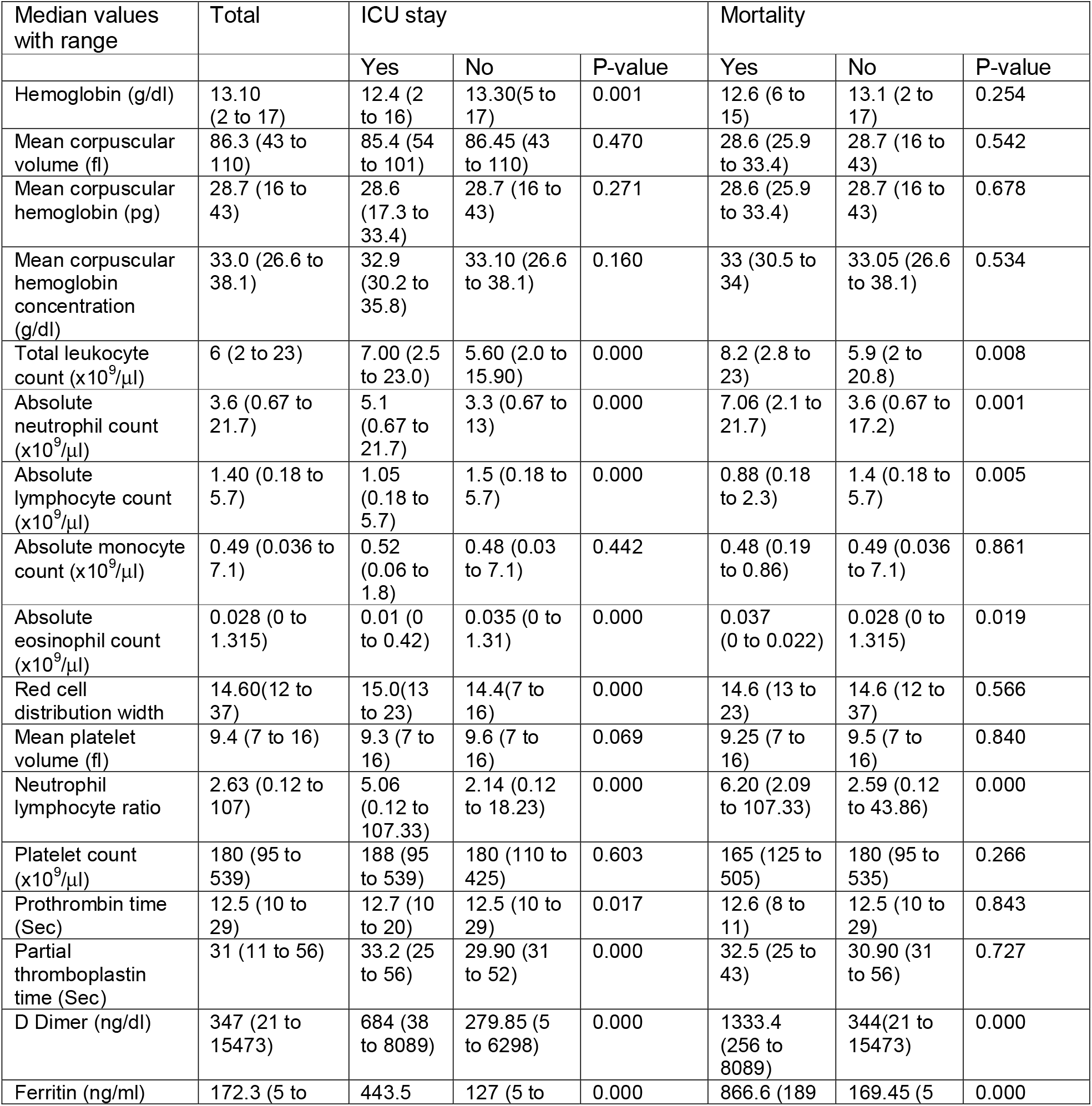
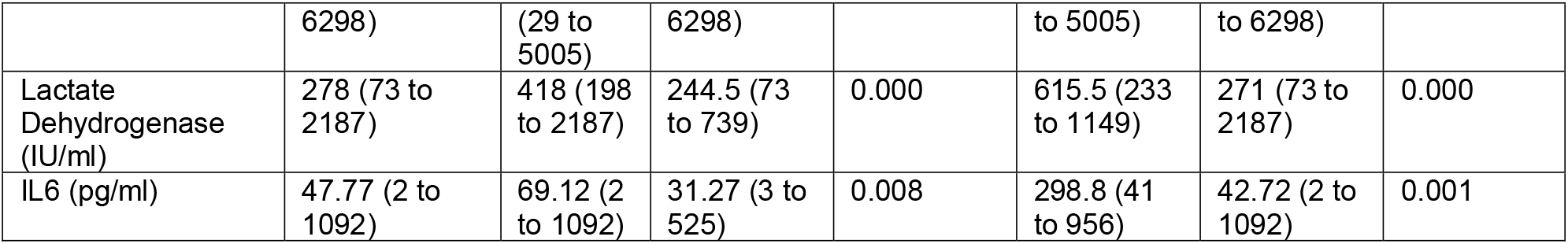
Hematological laboratory findings of COVID-19 patients on admission and association with outcomes.

**Table 3.** Hematological laboratory findings of COVID-19 patients on admission and association with outcomes.

|  | Total | ICU admissions |  |  | Mortality |  |  |
| --- | --- | --- | --- | --- | --- | --- | --- |
|  |  | Yes | No | P value | Yes | No | P value |
| Lymphopenia < $1 \times 10^9/\mu\text{l}$ | 107 | 56 | 51 | 0.000 | 9 | 98 | 0.003 |
| Lymphocyte count > $1 \times 10^9/\mu\text{l}$ | 329 | 69 | 260 | | 6 | 323 | |
| Eosinopenia < $0.02/( \times 10^9/\mu\text{l})$ | 195 | 78 | 117 | 0.000 | 11 | 184 | 0.033 |
| Eosinophil > $0.02/( \times 10^9/\mu\text{l})$ | 237 | 47 | 190 | | 4 | 233 | |
| Prolonged PT > 13 sec | 266 | 95 | 171 | 0.392 | 10 | 256 | 0.437 |
| Normal PT | 14 | 3 | 11 |  | 1 | 13 |  |
| Prolonged aPTT > 35 secs | 46 | 35 | 11 | 0.000 | 1 | 45 | 1.00 |
| Normal aPTT | 137 | 40 | 97 |  | 5 | 132 |  |
| Leukocytosis > $11( \times 10^9/\mu\text{l})$ | 36 | 25 | 11 | 0.000 | 5 | 31 | 0.000 |
| TLC < $11( \times 10^9/\mu\text{l})$ | 401 | 100 | 301 | | 10 | 391 | |
| Hb < 11 g/dL | 67 | 30 | 37 | 0.001 | 5 | 62 | 0.063 |
| Hb > 11g/dL | 370 | 95 | 275 |  | 10 | 360 |  |

**Table 4.**
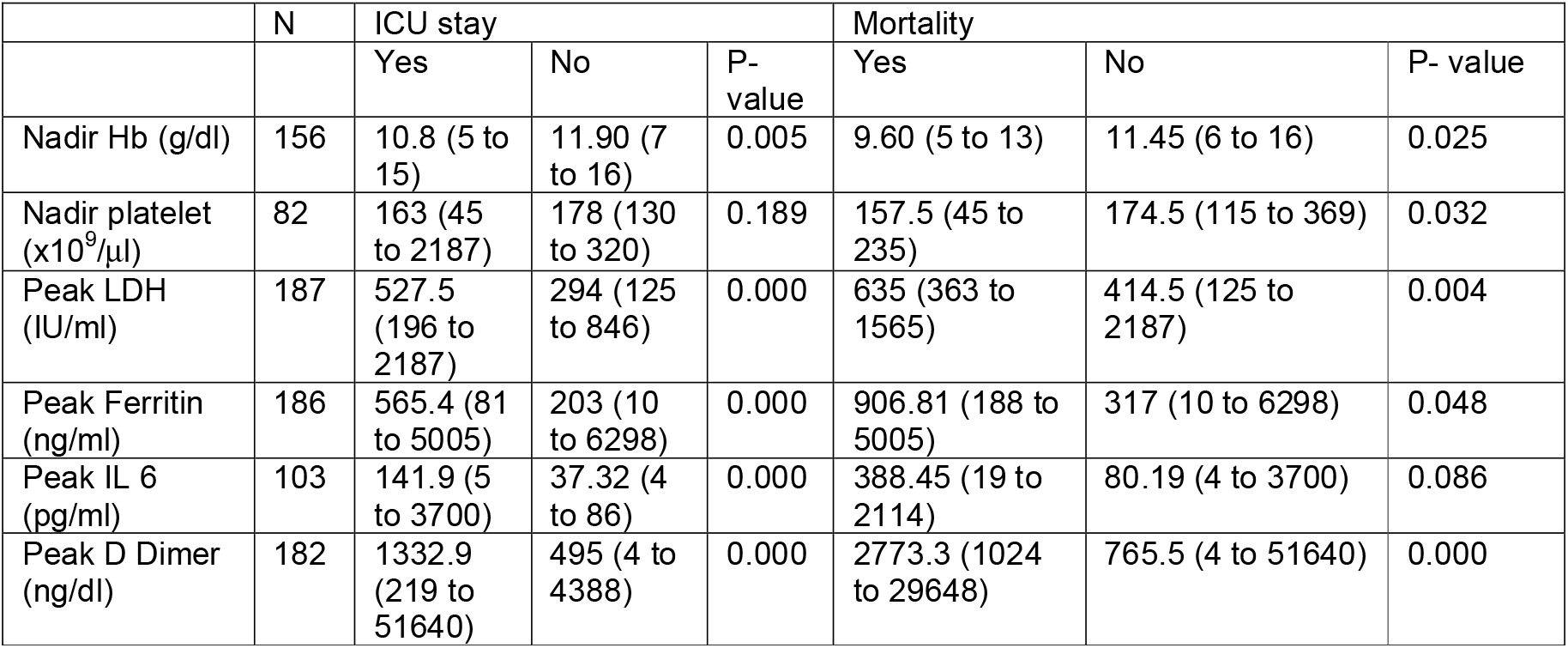
Peak or nadir hematological laboratory findings of COVID-19 patients on admission and association with outcomes.

Multivariate logistic regression analysis demonstrated that only serum ferritin and IL6 levels were independently associated with mortality. Ferritin cutoff ≥751 ng/ml and was able to identify 9/13 deaths and IL6 levels ≥64pg/ml was able to identify 8/9 deaths. When both cutoffs were used, all deaths were correctly identified. Ferritin (0.989) and IL-6 (0.985) had very high negative predictive value. These finding needs to be interpreted with caution in view of small number of events.

## Discussion

Covid-19 is a new illness and efforts are underway to identify simple prognostic markers that can be widely utilised. Focus on hematology of Covid has been surprisingly less. Complete blood count is a simple test which is performed for all patients, readily available at all healthcare facilities. Lymphocytes have an important function in viral immunity. Therefore, it is logical to think that patients with lymphopenia may be at higher risk of severity of illness and thereby mortality. We noted that patients with lymphopenia had poorer outcomes. Also, leukocytosis, even though present in minority of patients (8.2%) was associated with poorer outcome. The 2 variables combined leading to high NLR further increased the predictability of severity of illness or mortality.

In severe infections, neutrophils are often increased and activated with delayed apoptosis disorder[13]. The lymphopenia observed in patients with Covid-19 may be explained by their recruitment at inflamed sites and concomitant use of corticosteroids[9]. Neutrophilic leukocytosis along with lower lymphocyte and monocyte counts has been documented as an important finding in these patients[14]. This leads to altered NLR. On the other hand, Yang et al, reported leucopenia, lymphopenia and lower than normal range neutrophil counts and platelet counts[15]. Liu et al have associated high NLR to high mortality in these patients[8]. These differences may be due to different patient profiles with different severity. NLR, however, is a simple reliable ratio easily calculated by a routine blood count test.

Lower platelet count has been regarded as important predictor of mortality[3,10]. However, it is important to note that this finding is more of statistical significance and not of much clinical significance. Liao et al documented that median platelet counts were more than 100,000/cmm in all group of patients even though thrombocytopenia (<100,000/cmm) was seen in 48% of critical patients but degree of thrombocytopenia was not mentioned[3]. Another study identified lower platelet count in patients who dies vs those who survived (169 vs 213, p=0.009) but again this was not clinically relevant as all patients had normal platelet counts. Two other studies have demonstrated that thrombocytopenia is associated with mortality[15,16]. This is in contrast to our findings. In our study, only one patient had thrombocytopenia and baseline platelet count did not correlate with severity of illness or mortality. However, nadir platelet counts were associated with mortality (p=0.032). Mechanism of ow platelets in Covid-19 is largely consumptive in nature[6,17] even though immune thrombocytopenia have been reported in these patients[18].

Older age and presence of comorbidities continue to affect outcome negatively. A recent meta-analysis of 51,225 patients has highlighted age and comorbidity to be an important predictor of mortality[19]. Median age of patients who died was significantly higher in our study (65.1 years vs 46.5 years; p= 0.000). Presence of any comorbidity was significantly was associated with adverse outcome either for need for ICU transfer or mortality.

There have been attempts to identify other prognostic markers. In limited dataset of our patients, IL 6 levels correlated with adverse outcomes. A recent metanalysis conforms this finding where raised IL6 levels (with different cut-offs used in different studies) were independently associated with poor outcome[20]. Ferritin is an acute phase reactant and rises in host of infective, inflammatory situations. Similarily, high serum ferritin levels have been associated with severity of illness, comorbidities and adverse outcome[21]. Importance of high levels of these markers is of unpredictable importance. A recent randomised controlled trial has shown ineffectiveness of tocilizumab in reducing severity of illness or mortality in patients with Covid-19[22]. In our study, significant measurements associated with mortality were ferritin and IL6 levels as obtained by ROC curves. When serum, ferritin cut off ≥751 ng/ml and IL6 levels ≥64 pg/ml were combined then all deaths were correctly identified. Ferritin (0.989) and IL-6 (0.985) had very high negative predictive value. Even though the sensitivity and specificity was good with very high negative predictive values, but the predictivity was still low largely due to small number of patients who died.

## Limitations

This is a retrospective study. Therefore, complete data was not available for all patients. Covid-19 being a new disease, national guidelines for testing and need for hospitalisation/home isolation kept on changing. Many patients in this study were hospitalised who now in November 2020 are being managed as outpatients. This study largely focussed on baseline blood counts and did not follow the dynamic nature of blood count fluctuations which may happen during the course of illness. Patients have been treated differently and therapeutic agents used to treat may have affected peak or nadir values as highlighted in results. Study does not include peripheral smear findings. Mortality events were low. Data regarding predictors of mortality therefore needs to be interpreted with caution. For example lower platelets counts (within normal range) are associated with severity and risk of mortality. These limitations not withstanding we are able to identify simple prognostic markers.

## Conclusions

Peripheral blood counts at time of hospitalization is a simple tool to predict outcomes in patients admitted with Covid-19.

## Data Availability

All data produced in the present work are contained in the manuscript

